# Modeling COVID-19 Transmissions and Evaluation of Large Scale Social Restriction in Jakarta, Indonesia

**DOI:** 10.1101/2020.10.30.20222984

**Authors:** A. Hasan, Y. Nasution, H. Susanto, E.R.M. Putri, V.R. Tjahjono, D. Puspita, K. Sukandar, N. Nuraini, W. Widyastuti

## Abstract

This paper presents mathematical modeling and quantitative evaluation of Large Scale Social Restriction (LSSR) in Jakarta between 10 April and 4 June 2020. The special capital region of Jakarta is the only province among 34 provinces in Indonesia with an average Testing Positivity Rate (TPR) below 5% recommended by the World Health Organization (WHO). The transmission model is based on a discrete-time compartmental epidemiological model incorporating suspected cases. The quantitative evaluation is measured based on the estimation of the time-varying effective reproduction number (ℛ_*t*_). Our results show the LSSR has been successfully suppressed the spread of COVID-19 in Jakarta, which was indicated by ℛ_*t*_ < 1. However, once the LSSR was relaxed, the effective reproduction number increased significantly. The model is further used for short-term forecasting to mitigate the course of the pandemic.

## 1. Introduction

The government of Indonesia has been criticized by its medical experts for its approach to handle the COVID-19 outbreak. Indonesia is among countries with the lowest testing rates, which potentially masks the scale of its outbreak. The first COVID-19 cases in Indonesia were confirmed on March 2, 2020, with two citizens tested positive. The spread of the infection in Indonesia is marked by super-spreading events [1]. At the beginning of the outbreak, the government introduced two terms for people who have symptoms but have not had a test yet: Person under Observation (ODP) and Patient under Surveillance (PDP). The difference between ODP and PDP lies in the severity of the symptoms. PDPs have more severe symptoms that need to be hospitalized. According to the Indonesian Health Ministry, a person who has made contact with people who are positive with COVID-19 or has been traveled to COVID-19 epicenter within 14 days before the onset of the symptoms will be categorized as ODP or PDP. ODPs and PDPs must be self-isolated at home or hospital and monitored by the local health agencies until they have negative test results. At the onset of the outbreak, the number reached hundreds of thousands. Many of them died before getting their results and were not reported as fatality cases due to COVID-19. Lack of testing has become the main challenge in estimating the true scale of the outbreak.

Since 13 July 2020, the term ODP and PDP were no longer be used. Instead, The government of Indonesia introduced a new classification called “suspected cases” to address individuals who should be tested for COVID-19. According to the Indonesia Ministry of Health, a suspect is a patient who fulfills one of the following criteria: diagnosed with Upper Respiratory Tract Infection or URI (fever more than 38°C, and at least one symptom of respiratory illness like a cough or sore throat) and a history of travel or residence in a location reporting community transmission of COVID-19 disease during the 14 days before symptom onset, diagnosed with URI and having been in contact with a confirmed or probably COVID-19 in the last 14 days prior to symptom onset, or diagnosed with severe URI/pneumonia which requiring hospitalization and the absence of an alternative diagnosis that thoroughly explains the clinical presentation. In this paper, the number of suspected cases is obtained by combining ODP and PDP data. Figure 1 shows the number of suspected cases compared to active, recovered, and deceased cases.

**Figure 1:**
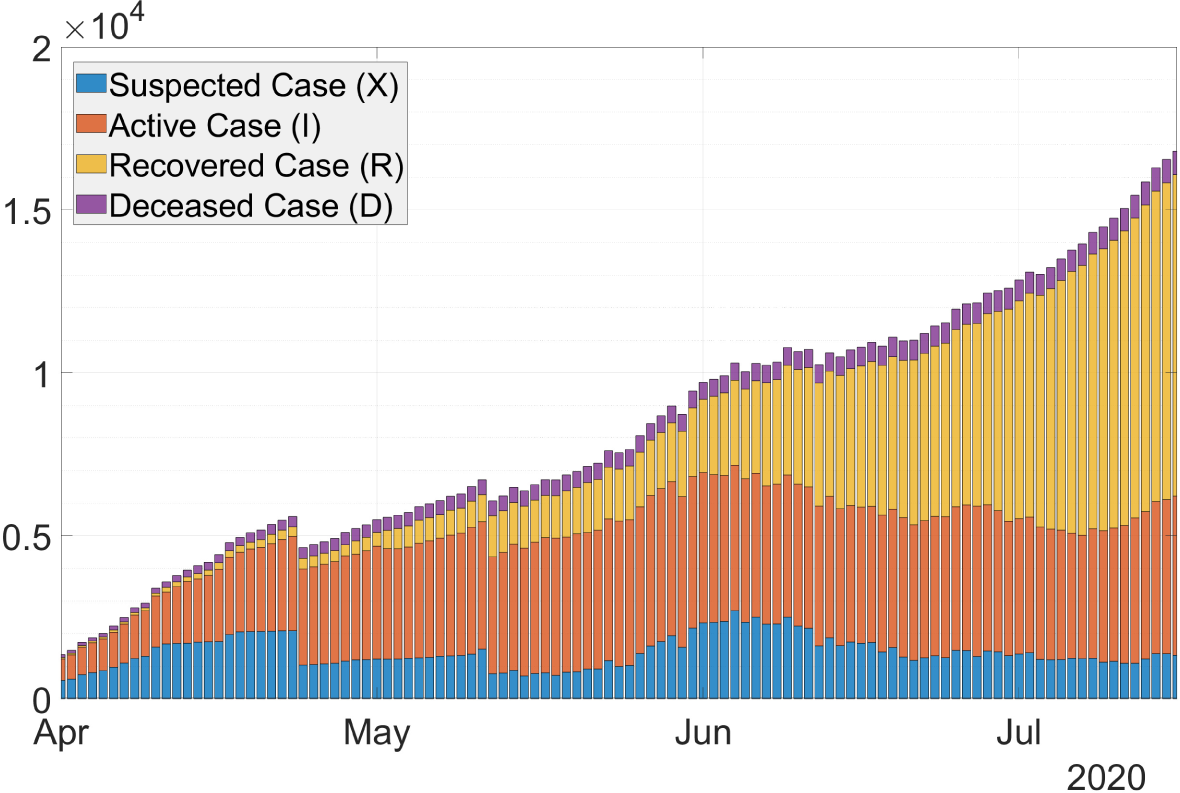
Stacked-bar of COVID-19 cases in Jakarta, Indonesia.

At the end of March, the cases soared to 1,200 cases, with 114 deaths. More than half of those cases and deaths came from Jakarta, more than any other provinces in Indonesia. Jakarta was indeed considered as the epicenter of the outbreak. On 31 March, the Indonesian government declared the outbreak as a national disaster. It announced stronger measures than social distancing to suppress the virus’s spread, which was called Large Scale Social Restrictions (LSSR). The LSSR allows regional governments to restrict the movement of people and goods within the regions.

The city of Jakarta became the first region to implement LSSR. The first stage of LSSR in Jakarta was implemented for two weeks, from 10 to 24 April 2020. During this period, schools, workplaces, public facilities, and places of worship were temporarily closed. Workers who are allowed to leave their houses would be required to wear face masks. LSSR restricted motorcycle taxis from carrying passengers and made it obligatory for hotels to accept people who are self-isolating. The LSSR has been extended twice. The first extension (LSSR II) was from 24 April to 22 May and the second extension (LSSR III) was from 23 May to 4 June. The measure was relaxed afterward, mainly because of economic concerns. The government of Jakarta announced the so-called LSSR transition to prepare for the transition to *a new normal*. During this transition period, many measures were relaxed. For example, 50% of workers could work in the office, and worship places are open to the public.

This paper is organized as follows. In Section 2, we present a mathematical model incorporating suspected cases to describe COVID-19 transmission in Jakarta. A method to estimate the time-varying effective reproduction number (ℛ_*t*_) based on the Extended Kalman filter (EKF) is discussed in Section 3. In Section 4, we discuss the evaluation of LSSR in Jakarta from 10 April until 4 June. A short-term forecast using the model is presented in Section 5. Finally, conclusions and recommendations are presented in Section 6.

## 2. Mathematical Modeling

In this paper, we propose a new compartmental epidemic model for COVID-19 transmissions in Jakarta. The motivation behind this new model is due to the fact that there are significant numbers of PDP and ODP (referred to as suspected cases) who show COVID-19 symptoms but have not been tested. Our idea is to include this data when estimating the time-varying reproduction number ℛ_*t*_. Our approach to estimate ℛ_*t*_ is based on Extended Kalman Filter (EKF) implemented on a discrete-time stochastic augmented compartmental model [2].

Figure 2 shows the model diagram. Let us denote *S, X, I, R*, and *D* respectively as susceptible, suspected, active/confirmed, recovered, and deceased compartment. The force of infection from compartment *S* will enter compartment *I* and *X* equally. The only difference is that when the testing result in compartment *X* is negative, the individual will go back to compartment *S*, whilst positive they will go to compartment *I*. Thus, we denote *ϵ* as the negative testing rate, while *κ* is the positive testing rate. Individuals from compartment *X* who died will go to compartment *D* at the rate of *ω*. An infected individual from compartment *I* has two outcomes, either to *R* or to *D*. Thus, *γ* and *δ* denote the recovery and death rate, respectively.

**Figure 2:**
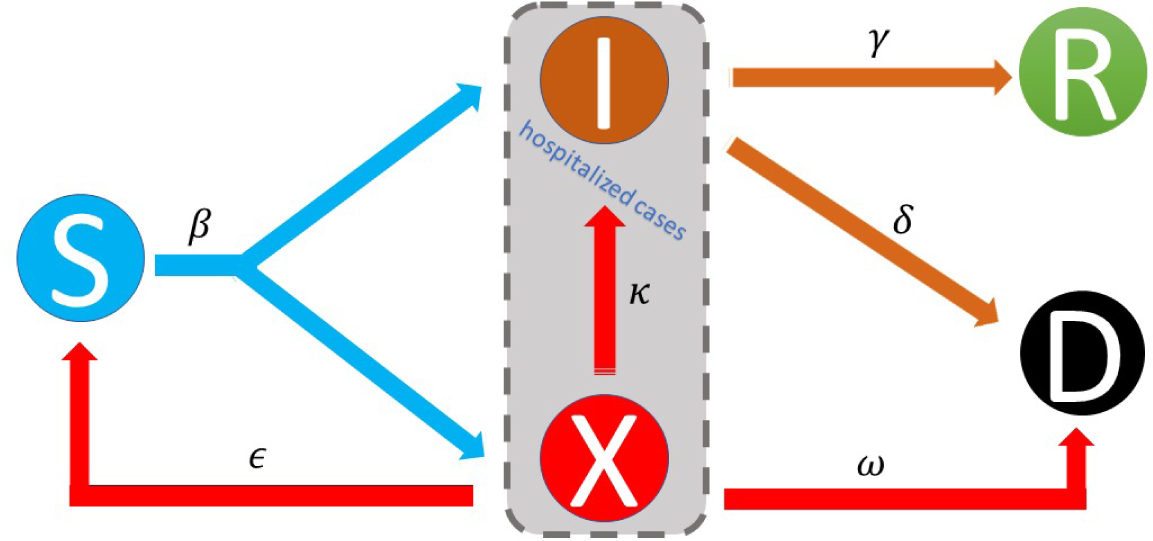
The SXIRD model is proposed to incorporate suspected cases of COVID-19 transmission in Jakarta.

### 2.1. Susceptible-Suspected-Infectious-Recovered-Deceased (SXIRD) model

The SXIRD model consists of five ordinary differential equations (ODEs) and is given by:

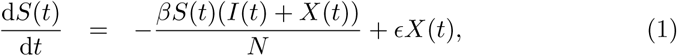

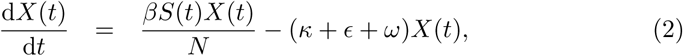

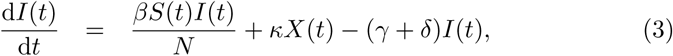

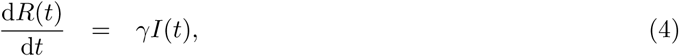

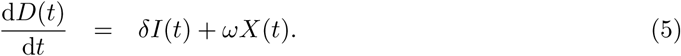

The system satisfies *S*(*t*) + *X*(*t*) + *I*(*t*) + *R*(*t*) + *D*(*t*) = *N*, where *N* denotes the total number of population. It can be shown using Picard-Lindelöf theorem [3] that (1)-(5) has a unique solution, i.e., for any initial condition *S*(0), *X*(0), *I*(0), *R*(0), *D*(0) ∈ ℝ, there exists *t*_0_ > 0 and continuously differentiable functions *S*(*t*), *X*(*t*), *I*(*t*), *R*(*t*), *D*(*t*) : [0, *t*_0_) → ℝ, such that the set of functions (*S*(*t*), *X*(*t*), *I*(*t*), *R*(*t*), *D*(*t*)) satisfies (1)-(5).

Let us assume that *S*(0), *X*(0), *I*(0), *R*(0), *D*(0) ≥ 0. The solution for (2) is given by:

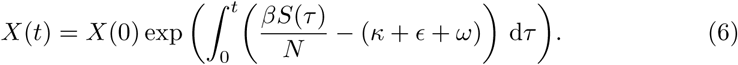

Thus, for *X*(0) ≥ 0 we have *X*(*t*) ≥ 0. Furthermore, the solution for (1) is given by:

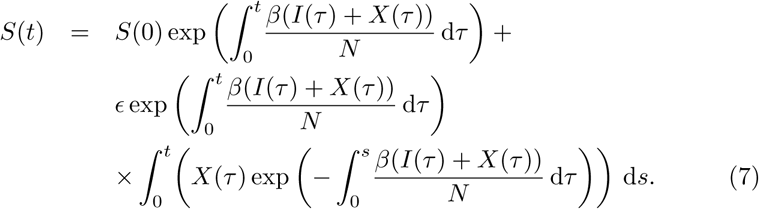

Since *X*(*t*) ≥ 0, then *S*(*t*) ≥ 0. Using similar approach, we can prove that *I*(*t*), *R*(*t*), *D*(*t*) ≥ 0. Thus the SXIRD is well-posed mathematically and biologically.

### 2.2. Time-varying reproduction number

Following Lemma 1 in [4], and [5], the basic reproduction number for the SXIRD model (1)-(5) is defined as the spectral radius of the next generation matrix and is given by:

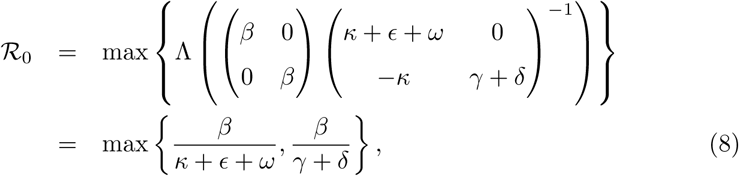

where Λ denotes the eigenvalue of the next generation matrix. Since in practice *β* = *β*(*t*) due to intervention, to take into account the decline number of susceptible individuals, the time-varying effective reproduction number is calculated using the following formula [6]:

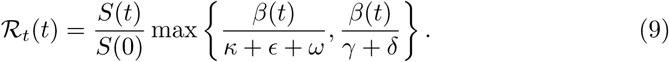

## 3. Estimation of the Time-Varying Effective Reproduction Number

In this section, we use EKF algorithm to estimate the time-varying effective reproduction number ℛ_*t*_(*t*). The EKF is an algorithm that provides estimates of unknown variables or parameters given measurements observed over time. To estimate ℛ_*t*_(*t*), the EKF is implemented to a discrete-time augmented compartmental SXIRD model.

### 3.1. Discrete-time augmented SXIRD model

Applying forward Euler method and augmented the infectious rate *β*(*t*) as a new variable, the discrete-time augmented SXIRD model is given by:

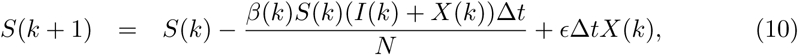

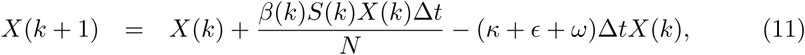

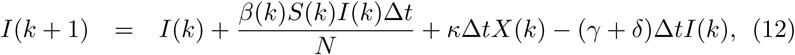

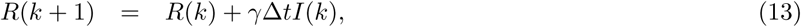

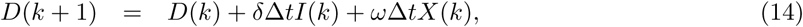

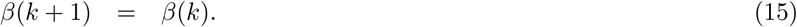

where Δ*t* denotes the time step. The last equation is obtained assuming the infection rate as a piece-wise continuous function with rare jumps. Indeed, the jumps are assumed to happen every one day when a new data is obtained.

### 3.2. Extended Kalman filter (EKF)

To simplify the presentation, we define an augmented state vector

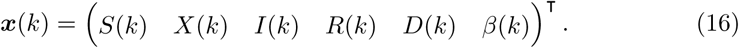

Thus, the discrete-time augmented SXIRD model (10)-(15) can be written as follows:

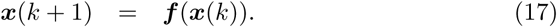

In order to use the EKF, we need to linearize the nonlinear model (17) about the posterior estimate of the previous state. Let us denote 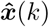 as the posterior estimate from the EKF in the previous step. Applying first-order Taylor series expansion to the nonlinear system (17) at 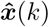, we obtain:

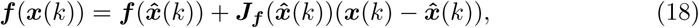

where 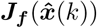 is the Jacobian matrix of ***f***, given by:

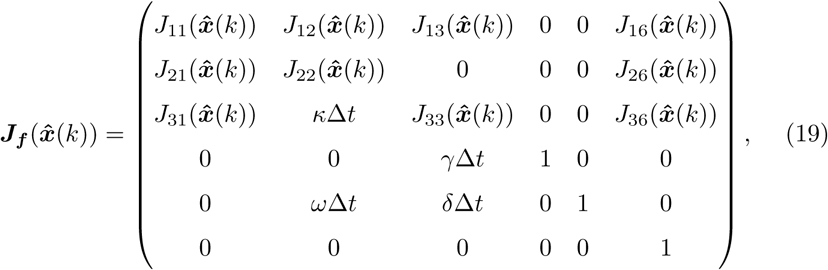

where

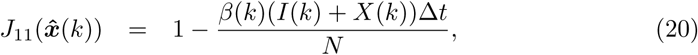

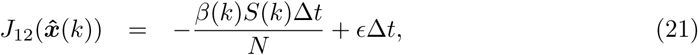

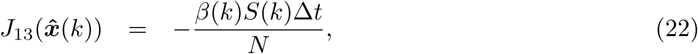

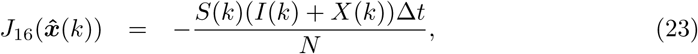

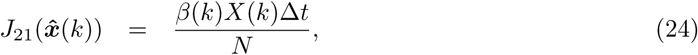

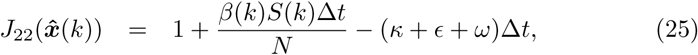

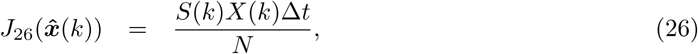

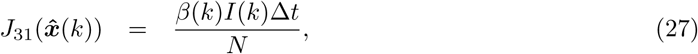

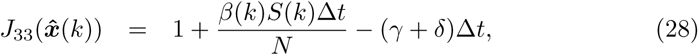

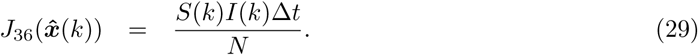

After we obtain the Jacobian matrix, we can use a standard EKF algorithm to estimate the state and the parameter [2]. The idea of EKF algorithm is to perform real-time data fitting on the suspected, active, recovered, and deceased cases. While performing the fitting, the algorithm produces the estimate of ℛ_*t*_ from (9). The EKF consists of two main steps: predict and update. In the prediction step, the method uses the model (17) to predict the state variable. The propagation of the state error covariance ***P*** is calculated based on the Jacobian matrix (19) and the error covariance of the model ***Q***_*F*_ (*k*). In this paper, ***Q***_*F*_ (*k*) is considered as a tuning parameter that minimize the error between the reported data and the estimated data. Let us denote 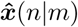 as the estimate of 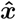 at time *n* given observations up to and including at time *m* ≤ *n*. The prediction step is given below:

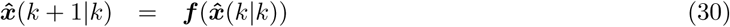

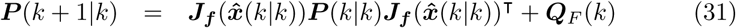

In the update step, first we compute the residual 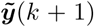. The residual is the difference between the reported data and the estimated data. The filter estimates the current data by multiplying the predicted state and the measurement/data matrix ***C***. Since ℛ_*t*_ is not measured, the data matrix is given by:

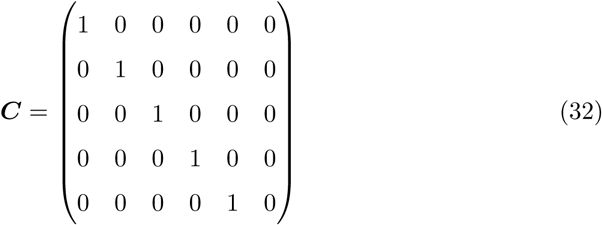

The residual is then multiplied by the Kalman gain ***K***(*k* + 1|*k*) to provide the estimation of the state variable 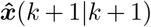. The update step is given below:

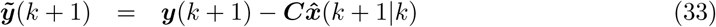

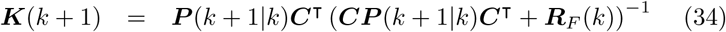

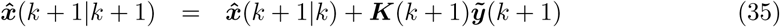

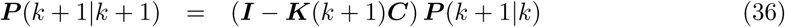

Here, ***R***_*F*_ (*k*) is also considered as a tuning parameter that minimize the error between the reported data and the estimated data. Figure 3 shows the results of EKF for real-time data fitting of COVID-19 cases in Jakarta. We can observe that the EKF estimates the number of cases accurately.

**Figure 3:**
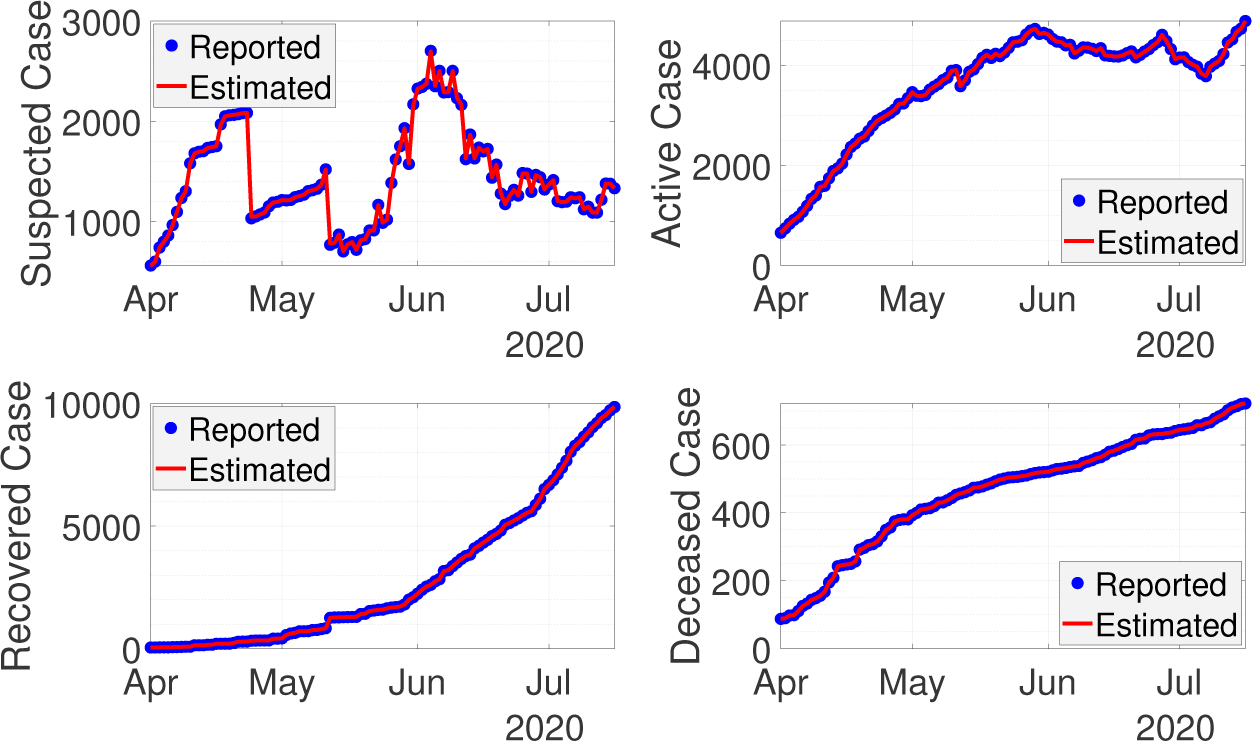
Real-time data fitting using EKF for COVID-19 cases in Jakarta.

## 4. Evaluation of the Large Scale Social Restriction (LSSR)

The local government of Jakarta used the time-varying effective reproduction number ℛ_*t*_ as an indicator to assess the duration of LSSR. The estimated ℛ_*t*_ was obtained from daily confirmed cases and without considering the number of suspected cases. In this section, we use the estimated value of ℛ_*t*_ incorporating the number of suspected cases to evaluate the LSSR in Jakarta. We first evaluate the difference between the estimated ℛ_*t*_ involving suspected cases and the one without suspected cases. In our calculation, we assume the infectious time *T*_*i*_ is 12 days with standard deviation of 3 days. The recovery rate *γ* and the death rate *δ* are given by 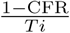 and 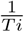, respectively, where CFR is the Case-Fatality-Rate. The negative testing rate *ϵ* is assumed equal to the removed rate, i.e., *ϵ* = *γ* + *δ*, while the positive testing rate *κ* is 20%. MATLAB implementation can be found here: https://github.com/agusisma/CovidJakarta. Figure 4 shows that in general the estimated ℛ_*t*_ with suspected cases included is slightly different from the one without suspected cases. Due to a lower number of suspected cases in Jakarta than other provinces, such as West Java province (see[7]), the estimation of ℛ_*t*_ in Jakarta gives almost similar results. From the third week of May until the first week of June 2020, however, there was a significant jump in the number of suspected cases. In this period, the estimated ℛ_*t*_ with suspected cases shows higher value. As the evaluation of LSSR is based on the estimated value of ℛ_*t*_, it is important to include the suspected cases in the model when the number of cases is high.

**Figure 4:**
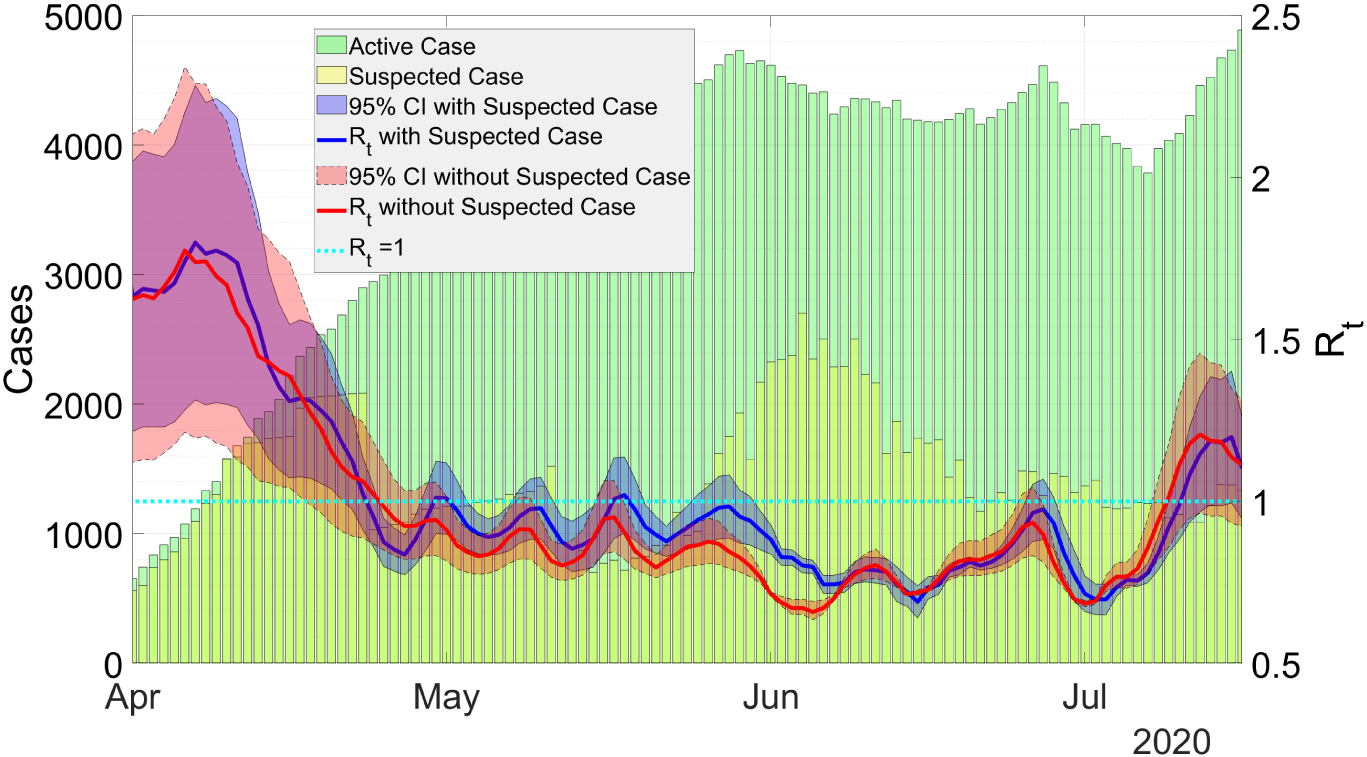
Estimation of ℛ_*t*_ with and without suspected cases.

In Figure 5, we plotted the normalized ℛ_*t*_ and the period of LSSR in Jakarta. The effect of LSSR can be shown in the next two weeks after the policy is applied because the symptoms of COVID-19 may appear 2-14 days after exposure [8]. During the first period of LSSR, starting from 10 April 2020, the estimated value of ℛ_*t*_ for both models are decreasing. It may happen as the effect of several policies of local government, such as social distancing, wearing mask, and work from home [9]. During the second period of LSSR until early LSSR transition period, the estimate value of ℛ_*t*_ continues decreasing and subsequently the value is less than 1. Consecutive decreasing of ℛ_*t*_ during LSSR I-III, indicates that the policy was applied successfully.

**Figure 5:**
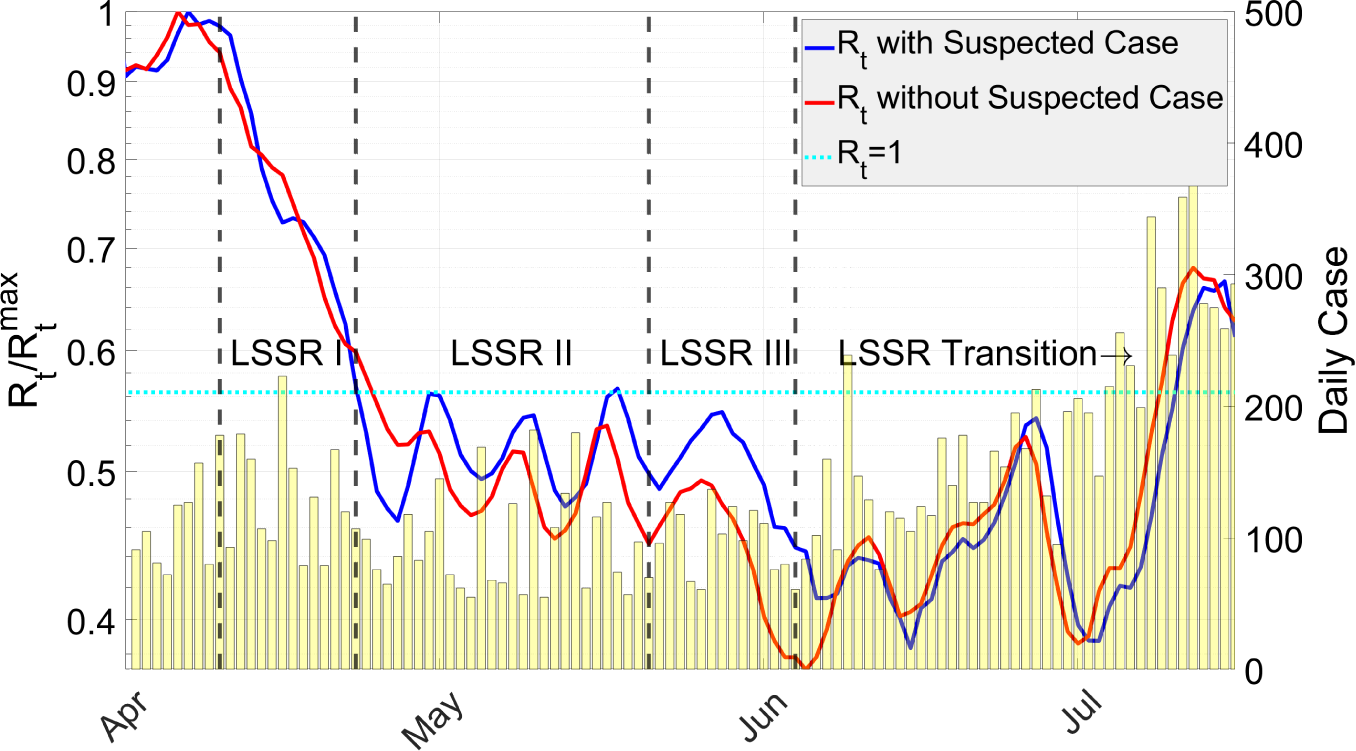
Mean of 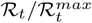 plotted in semi-log scale.

As ℛ_*t*_ < 1 for several weeks during the LSSR transition, the restrictions were relaxed gradually for economic reasons [10]. Unfortunately, easing the policy restrictions seems to result in the increase of value of ℛ_*t*_ > 1 in just a short time period. As a consequence, a more tightened policy should be considered by the local government of Jakarta to avoid the second wave of the outbreak. In order to apply a proper LSSR policy based on the estimated value of ℛ_*t*_, forecasts on the number of active, recovered, and deceased cases are required and are given in the next section.

## 5. Short-term Forecast

In this section, we provide short-term forecasts (60-day) on the number of COVID-19 cases. The forecasts are conducted in two presentations. First, by assuming the current measures are continued, a short-term forecast for the total number of cases with a 95% confidence interval is shown in blue-dashed line in Figure 6. The forecast is done by calculating the total case from (10)-(14). The actual total number of cases is shown in red-dashed line. From this figure, we can observe that our forecast provide a reasonable result. The longer the forecast, the wider the confidence interval. Furthermore, the forecast is conducted using three different scenarios. Scenario 1 represents the situation assuming LSSR is completely stopped and all activities go back to normal. This scenario is applied within the next two months and the result of the forecast are presented by the yellow-dashed lines in Figure 7. We can observe a sharp increase in the number of active as well as deceased cases. Scenario 2 represents the situation assuming current measure are continued and the forecasting results for the next two months are shown by the blue-dashed lines in Figure 7. Remark that, the current measures (per September 2020) taken by the local government of Jakarta is LSSR transition, which is a relaxed version of the original LSSR I-III. In this scenario, the number of infected individuals are increased. Scenario 3 is applied by assuming that the current measure is tightened. This scenario can be considered as the implementation of the original LSSR. The forecasting results are shown by the green-dashed lines in Figure 7.

**Figure 6:**
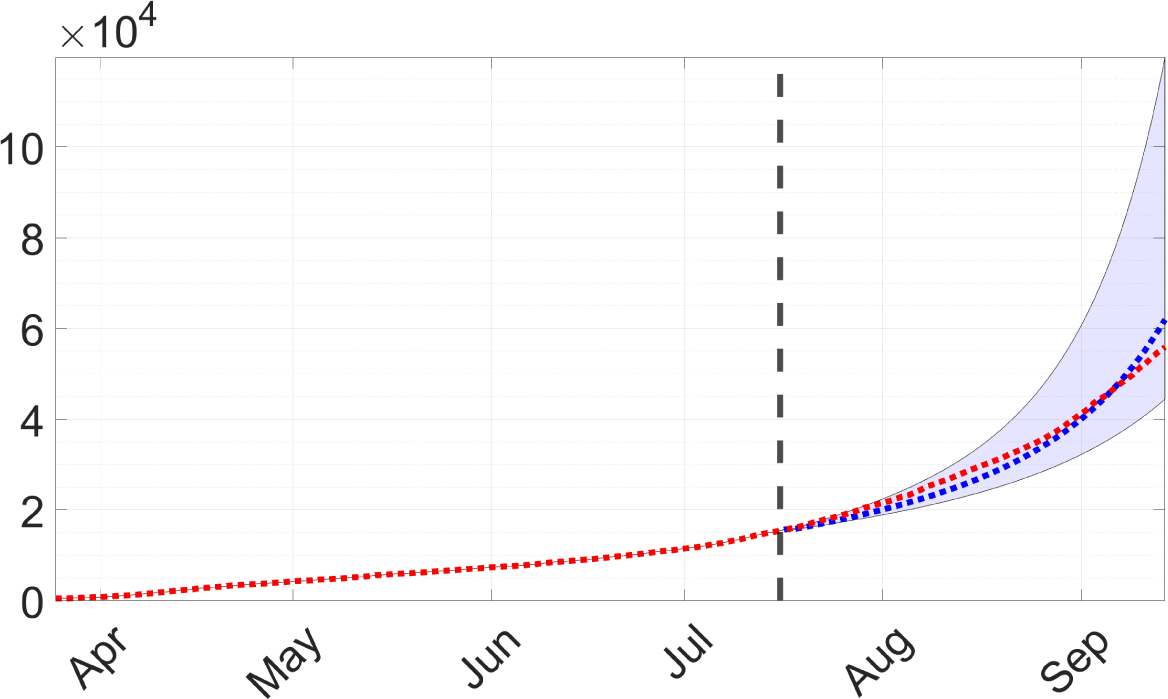
Short-term forecast for the total number of COVID-19 case. The red-dashed line is the actual total number of reported cases. The blue-dashed line is the projection from our approach.

**Figure 7:**
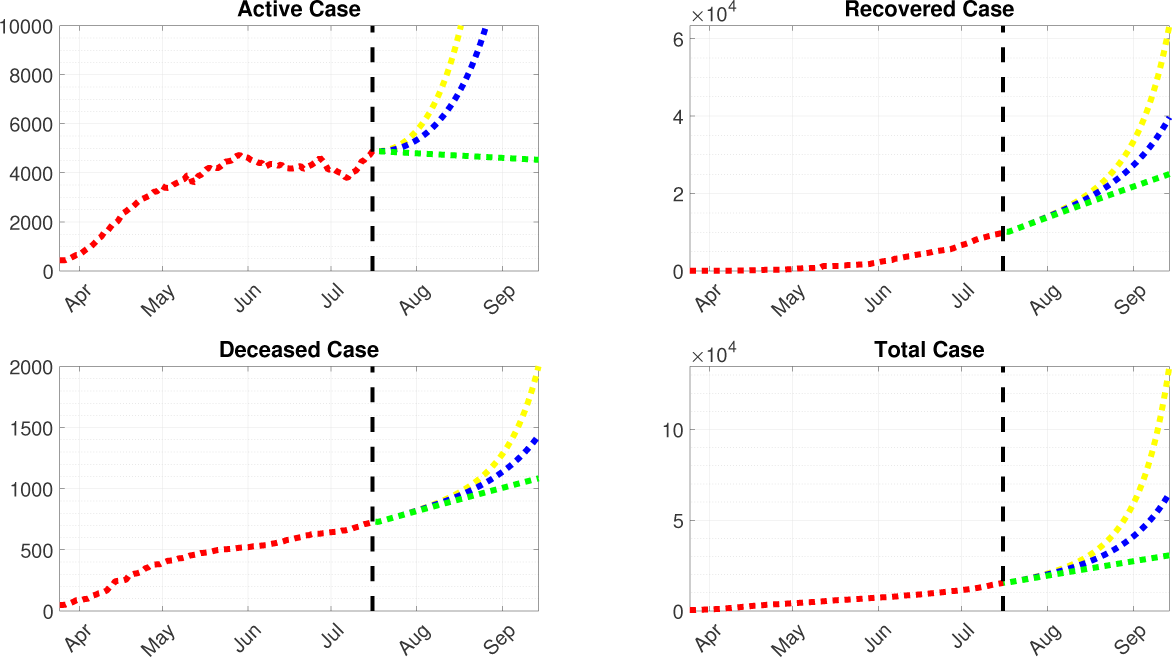
Forecast with different scenarios.

The results show that the number of active cases by Scenario 1 is significantly increased within the next two months, whereas Scenario 2 and Scenario 3 may result in a slight and an instant decrease on the number of cases. The number of deceased and recovered cases remain increasing for three scenarios during the first month. However, at the second month it shows different slopes. Among these scenarios, only when the current measures are tightened we can see declines in the number of active, deceased, and total cases.

## 6. Conclusions and Recommendations

We have presented a new model for COVID-19 transmission in Jakarta, Indonesia. The model incorporated the number of suspected cases and is used to evaluate the effectiveness of the LSSR and to forecast the number of COVID-19 cases under different scenarios. We conclude that the LSSR, which was implemented from 10 April until 4 June 2020, has been successfully reduced the transmission of COVID-19 in Jakarta. Looking at the development of cases after measures under LSSR are lifted, we recommend enacting the measures back to the original LSSR. Furthermore, we recommend the local government of Jakarta to increase the number of testing until the average Testing Positivity Rate (TPR) is below 5% again.

## Data Availability

Data are available and are given in the below link.

https://github.com/agusisma/CovidJakarta

